# Data-Driven Leisure-Time Physical Activity Trajectories of Over 46 Years and Their Associations with Cognition in Nonagenarians – A Cohort Study

**DOI:** 10.1101/2025.09.24.25336548

**Authors:** Paula Iso-Markku, Gabin Drouard, Vahid Farrahi, Anni Varjonen, Henri Vähä-Ypyä, Tommi Vasankari, Jaakko Kaprio, Eero Vuoksimaa, Sari Aaltonen

## Abstract

Based on short follow-up studies, greater leisure-time physical activity (LTPA) has been associated with better cognition in old age, but more longitudinal studies are needed. Our aim was to identify long-term LTPA trajectories from midlife to late old age and examine whether these trajectories are associated with nonagenarians’ cognition.

In total, 125 participants from the NONAGINTA – Memory and Health in Nonagenarians study were included. The participants responded to health surveys of the older Finnish Twin Cohort study including LTPA at the mean ages of 45, 52, 59 and 91 years. Cognition was assessed at the mean age of 91 years (standard deviation 1.54) via telephone interview (global cognitive function, episodic memory, and semantic fluency). We identified LTPA trajectories with K-means clustering for longitudinal data and we used generalized estimating equations models to investigate differences in cognition among the LTPA trajectories. Covariates included age, sex, and education.

We found three LTPA trajectories from midlife to nonagenarian age. The largest proportion of participants belonged to the *Constant low* trajectory (52%), characterized by a stable low level of physical activity throughout the follow-up. Two other trajectories were *Starting low and increasing* (25%) and *Starting high and decreasing* (23%). Cognitive measures at the mean age of 91 did not differ among the LTPA trajectories.

To conclude, we found three different trajectories of LTPA from midlife to late old age within a period of 46 years. There were no significant differences in cognition by distinct LTPA trajectories when age, sex, and education were considered.

## Introduction

Modifiable lifestyles have great potential in the prevention of dementia.^1^ In previous meta-analyses, physical activity has been associated with less cognitive decline in late life and a lower incidence of dementia.^2–3^ Causal evidence on the topic is lacking,^4^ but the challenges of randomized controlled trials remain to be sufficient compliance and sufficient length, especially when considering a disease with an exceptionally long pre-clinical phase. Observational studies can provide long-term or even life-course information on the possible protective factors of dementia, while avoiding the problem of poor compliance.

To date, most observational studies published on the association between physical activity and dementia or cognitive decline rely on physical activity measurements at a single time point.^2,3^ However, examining the association between longitudinal physical activity patterns and cognition would provide a more in-depth perspective into this association since the magnitude, rate, and timing of physical activity changes can then be considered. So far, only a few existing studies on physical activity and cognitive functioning have utilized longitudinal physical activity trajectory study designs. For example, Sabia et al.^5^ found that physical activity trajectories start to decline nine years before dementia diagnosis. This may be accounted for by reverse causation, i.e the effect of subclinical disease on the ability to exercise. Previous studies on longitudinal physical activity trajectories have also shown that those participants whose cognitive performance declines at ages 60–75 most likely belong to the low physical activity trajectories^6,7^ or that physical activity trajectories with a declining trend precede^8^ or are concurrent with cognitive decline.^9,10^ A recent study showed that it is not individuals who belonged to the highest or lowest physical activity trajectory but rather to the most stable physical activity trajectory that perform best cognitively.^11^ Additionally, the same authors have shown that the strength of the association between physical activity and cognition seems to weaken with aging.^11^ However, it must be noted that three of these abovementioned studies are from the same study cohort (i.e., the China Health and Retirement Longitudinal Study).^8,9,11^ In a study cohort of individuals under age 45, consistently high physical activity trajectories were shown to be associated with only one cognitive test: that is, better verbal fluency out of eight cognitive tests altogether.^12^

The participants of most previous studies investigating the association between physical activity trajectories and cognition have been older adults aged 70 to 80 years at the time of cognition assessment (see eFigure 1). Nevertheless, older adults aged 80 years and over are a rapidly growing age group – predicted to triple between 2015 and 2050.^13^ Even among older adults aged 85 years and over, nonagenarians are a unique group. Cross-sectional studies show that cognitively better-performing individuals aged 85 years and over seem to be physically more active.^14^ Only a few studies have explored the association of preceding physical activity and cognition among nonagenarians, with all population-based studies showing no association.^15–17^ Only the one volunteer-based study on the subject found a significant association between high physical activity levels and better subsequent cognition in nonagenarian women.^18^ However, to our knowledge, no nonagenarian study has investigated the association between physical activity and cognition, considering physical activity behavior from several time points over multiple decades.

Our aim was to identify 46-year leisure-time physical activity (LTPA) trajectories based on self-reports from midlife to the tenth decade of life and examine whether cognition in nonagenarians is associated with the LTPA trajectories identified. We used data-driven methods that enable finding patterns within the data without any influence of a priori assumptions. LTPA denotes commuting and other LTPA in this study in contrast to occupational physical activity that has differential association with health than LTPA.^3^

## Methods

The participants of the current study were from the longitudinal population-based dataset of the Older Finnish Twin Cohort study launched in 1975.^19^ The cohort consists of twins from same-sexed pairs born in Finland before 1958. The participants have responded to the mailed health and behavior surveys in 1975, 1981, and 1990 (response rates 77–89%). In the current study, we used data from all three surveys.. Health survey and cognitive interview data from the NONAGINTA – Memory and Health in Nonagenarians (response rate 27%) sub-study of all twins who reached 90 years of age was conducted in 2020–2024 was defined as the fourth and latest follow-up time point.^20^

To create longitudinal LTPA trajectories from midlife to the tenth decade of life, we included those twins in the study who had LTPA data and were between ages 42–51 (mean age 45.2 years) at baseline. In 1981 and 1990, the participants were at the mean ages of 51.6 years (age range 48–58) and 59.2 years (age range 57–60), respectively. At the last follow-up time point, the participating twins had reached the mean age of 91.2 years (age range 90–97). Hereafter, the mean ages are referred to as 45, 52, 59 and 91. In total, we had LTPA data from at least two time points available from 125 twin individuals (65% women), including 14 complete twin pairs: 107 individuals at age 45, 112 individuals at age 52, 66 individuals at age 59, and 125 individuals at age 91. Most participants had LTPA data from 3 or 4 time points (106 participants, 85%). Global cognitive functioning data were available for 62 individuals, while episodic memory (immediate and delayed recall) and semantic fluency data were available for 80 individuals.

### Leisure-time physical activity

Participants reported their LTPA behavior at all survey time points by using the structured and validated questions on commuting and LTPA.^21–22^ In these questions, participants reported their monthly frequency, duration, and intensity of LTPA, including active commuting to and from work at mean ages 45, 52, and 59 (we assumed there is no commuting activity in nonagenarians). The LTPA questions were included in all survey questionnaires in the same form, except at mean age 59 in 1990 when only one question was used. This question measures combined information on the frequency, duration and intensity of LTPA, including commuting activity. The exact questions and response options for the questions are given in the Supplemental Material. The overall LTPA level that was used to create LTPA trajectories from midlife to the tenth decade of life was quantified as metabolic equivalent of task (MET) hours expended per day. The calculation of MET hours per day is described in detail in the Supplemental Materials and Methods.

Accelerometer-measured physical activity was assessed in a subsample of 91-year-old participants (N=36). These participants used a hip-worn tri-axial accelerometer for seven days to monitor their daily physical activity (see Supplemental Materials and Methods and Aaltonen et al.^20^).

### Cognitive measures

We used four different cognitive measures at age 91, all based on telephone interviews. We measured global cognitive function with the modified version of the validated Telephone Interview for Cognitive Status (TICS-m3) with additional modification to include three learning trials in the word list learning task.^23^ The TICS-m3 is a short, 15-item test of cognitive functioning of 4 domains: orientation, memory, attention, and language (score 0 to 70). Episodic memory was measured with a 10-word list learning task as a part of the TICS-m3. We used the total number of words recalled in three learning trials (total score ranges from 0 to 30) as a measure of immediate recall, and the number of words recalled after a delay of about 5 minutes as delayed recall (total score ranges from 0 to 10).^24^ Semantic fluency was assessed by a one-minute animal naming task. The telephone interviews were carried out by a trained research nurse. As a sensitivity analysis, an older version of the TICS-m was also analyzed.^25^ The TICS-m only has one learning trial but is otherwise like the TICS-m3.

### Covariates

The sex recorded at birth and date of birth were received from the Central Population Registry of Finland. We used the highest educational attainment (years schooling) the participants had reported in 1975, 1981, or as nonagenarians as a measure of education.^23,26^

### Ethics

The data collection was approved by the ethics committees of the Hjelt Institute, University of Helsinki and the Helsinki and Uusimaa Hospital District, Finland. The latest ethical approval was given by the Ethics Committee of the Hospital District of Helsinki and Uusimaa for the NONAGINTA study protocol on May 8, 2020 and December 16, 2020 (the latest follow-up time point in the current study). All study methods were carried out in accordance with the approved guidelines and the Helsinki Declaration. All participants provided their written informed consents for the study.

### Statistical analyses

#### Data-driven longitudinal leisure-time physical activity trajectories analysis

We used K-means cluster modeling for longitudinal data (KmL) to create and identify LTPA trajectories over a 46-year follow-up period with a total of up to 4 repeated measurements of physical activity.^27^ KmL is an enhanced version of the widely used K-means clustering algorithm, specifically designed to analyze and group longitudinal data consisting of repeated observations over time. Compared to the traditional K-means clustering algorithm, KmL is particularly well-suited for identifying clusters in longitudinal datasets with repeated measurements that are measured across multiple time points.

Prior to trajectory analysis, we excluded 2 outliers with over 15 MET-hours per day (corresponds to about a 1.5-hour run at a pace of 10 kilometers per hour) when participants were at mean age 59 in 1990 and normalized the data using the min-max normalization method. We allowed for a maximum of two missing time points. We specified the analysis to find from 2 to 6 LTPA trajectories with 500 permutations for each number of groups. Because different clustering validation measures have different weaknesses^28^ and no singular index is superior,^29^ we relied on several different clustering indices. The optimum number of trajectories was chosen based on Calinski–Harabatz (with three versions), Ray–Turi and Davies–Bouldin indices, as well as the sizes of the identified subgroups and the clinical relevance and stability of the LTPA trajectories. The number of clusters that yields the maximum index is preferred over other solutions. We prioritized the Davies–Bouldin index over the Calinski–Harabasz index because it has been shown to provide more reliable results.^29^ KmL for longitudinal data was performed with the KmL package in R version 4.3.3 and RStudio version 2023.12.1 Build 402.^27^

### Longitudinal leisure-time physical activity trajectory characteristics and their associations with cognition

When LTPA trajectories were identified, we continued analyses by investigating the associations between LTPA trajectory membership and cognition characteristics using generalized estimating equations (GEE) models (the ‘geepack’ package in RStudio, R version 1.3.9). GEE models account for the non-independence of observations arising from family relatedness. We conducted two sets of analyses. In the first set of analyses, we compared individuals in each trajectory to those in the other two trajectories, while in the second set of analyses, we involved pairwise comparisons between the trajectories. In both sets of analyses, we modeled LTPA trajectories as outcomes and cognition characteristics as independent variables. Continuous cognitive measures were used as such in the analyses. Both sets of analyses were first conducted with sex and age as covariates, and then with sex, age, and education as covariates. We tested the null hypothesis that the coefficients of the cognitive variables were equal to zero and reported odds ratios along with their 95% confidence intervals and p-values. P-values were reported both as nominal and corrected for multiple testing within each set of analyses using the Benjamini–Hochberg procedure.

### Longitudinal leisure-time physical activity trajectory characteristics and their associations with nonagenarians’ accelerometer-measured physical activity

To validate self-reported LTPA, we investigated the associations between LTPA trajectory membership and accelerometer-measured physical activity data in a subsample of nonagenarian participants^20^ using GEE models. We compared individuals in each trajectory to individuals in the two other trajectories, and sex and age were used as covariates.

### Sensitivity analysis

As sensitivity analyses, we compared the differences in study participants’ characteristics (sex and the length of education) between those who did and did not participate in the cognition assessment, using a t-test for continuous variables and χ^2^ test for categorical variables (adjusted for clustered twin data) (Stata statistical software version 19.5, StataCorp, College Station, Texas, USA). Furthermore, we did a sensitivity analysis to confirm the LTPA trajectories obtained were not dependent on missing values (R version 4.3.3 and RStudio version 2023.12.1 Build 402). We further compared the differences in the length of education between the participants who belonged to different LTPA trajectories using analysis of variance (ANOVA) (Stata statistical software version 19.5, StataCorp, College Station, Texas, USA).

## Results

The mean levels of LTPA among the full study cohort were 2.1 (standard deviation (SD) 1.8) MET-hours per day at age 45, 2.3 (SD 1.6) MET-hours per day at age 52, 2.4 (SD 1.9) MET-hours per day at age 59, and 1.6 (SD 1.7) MET-hours per day at age 91.

### Longitudinal leisure-time physical activity trajectories

Regarding longitudinal LTPA trajectories, we present clustering validation indices for the final solution and six examples of clustering solutions in Supplemental Table S1 and Supplemental Figures S2–3. The clustering validation indices were not in agreement, as can be seen in Supplemental Figure S2 and Supplemental Table S1. Most validation indices suggested good clustering with 3 or 4 groups, while some validation indices were poor when there were 2, 5, or 6 groups. Because of our small sample size and the better stability of 3 LTPA trajectories, we chose a clustering solution with 3 trajectories among the options of 3 or 4 trajectories. This decision also follows the general statistical principle of parsimony. From the 500 permutations with 3 trajectories, we chose the second redrawing with the more balanced sizes of the groups and lower Davies–Bouldin index than in the first redrawing (Supplemental Table S1 and Supplemental Figure S2). The mean trajectories for six different clustering solutions (from 3000) are presented in Supplemental Figure S3. After repeating the analysis excluding the participants with missing values, the shapes of the 3 LTPA trajectories remained similar.

We labeled the three distinct longitudinal LTPA trajectories identified by their unique elements as: *Constant low* (52.0%), *Starting low and increasing* (24.8%), and *Starting high and decreasing* (23.2%) (Table 1, Figure 1). The largest proportion of participants belonged to the *Constant low* trajectory. This trajectory was characterized by low LTPA throughout the follow-up, with a trend of a slight decrease: the mean leisure-time MET-hours per day were 1.2 (SD 1.0), 1.6 (SD 0.9), 1.2 (SD 0.9), and 0.7 (SD 0.7) at the mean ages of 45, 52, 59, and 91 years, respectively. These values approximately correspond to a daily 30-minute walk at a slow pace in midlife and a daily 15-minute walk at a slow pace at the mean age of 91.

**Figure 1.**
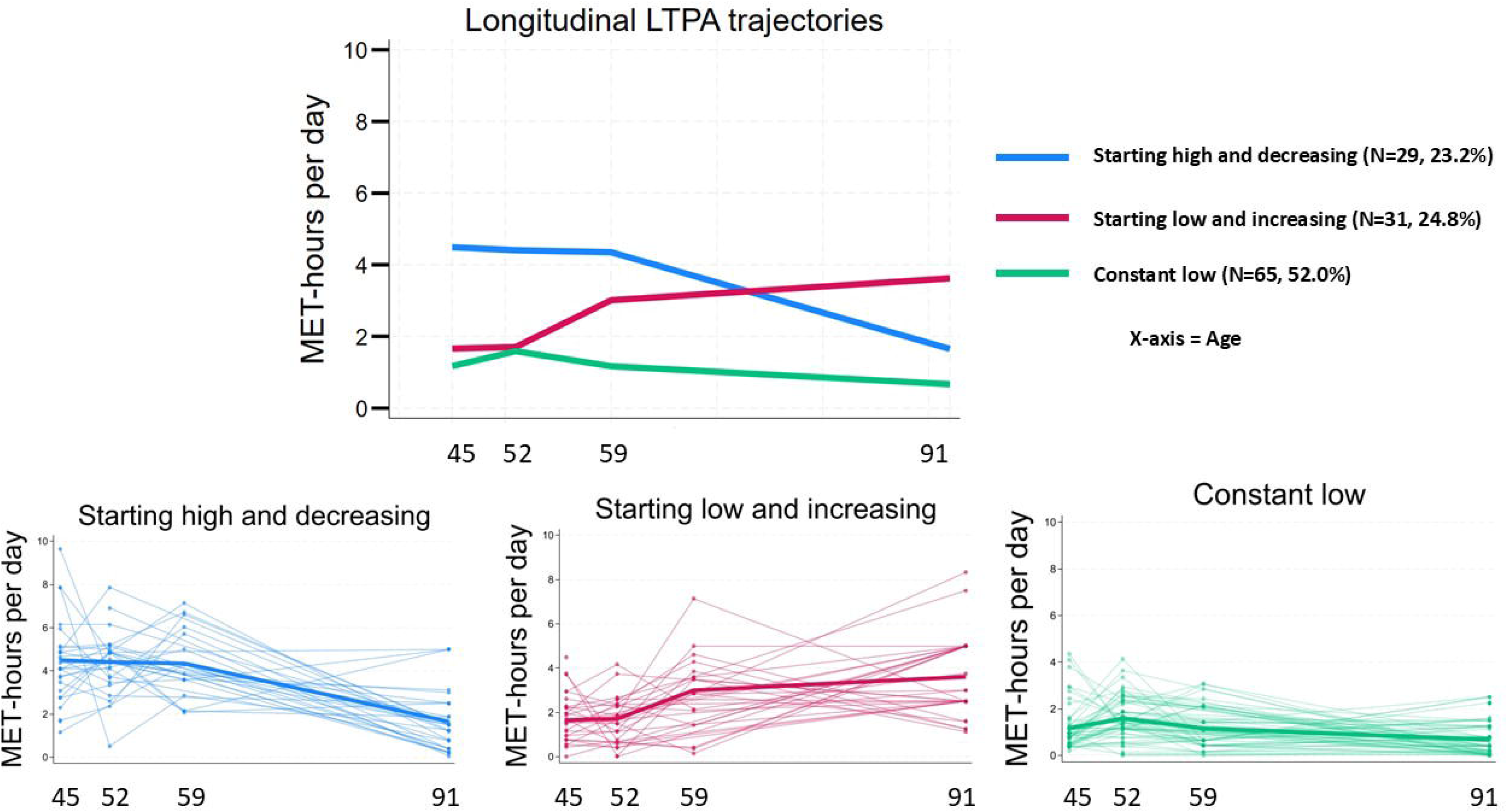
Leisure-time physical activity trajectories from midlife to the tenth decade of life with the spaghetti plots of individual trajectories. LTPA=leisure-time physical activity.

**Table 1.** Characteristics of the study cohort by longitudinal leisure-time physical activity trajectories.

| Characteristic variables | LTPA trajectories |  |  |
| --- | --- | --- | --- |
|  | Constant Low | Starting High and Decreasing | Starting Low and Increasing |
| **Number of study participants** (n, %) | 65 (52.0) | 29 (23.2) | 31 (24.8) |
| **Sex** |  |  |  |
| Women (n, %) | 45 (69.2) | 18 (62.1) | 18 (58.1) |
| Men (n, %) | 20 (30.8) | 11 (37.9) | 13 (41.9) |
| **Years of education** (mean, SD)* | 7.5 (3.1) | 10.2 (4.9) | 8.4 (4.2) |
| **MET-hours per day age 45** (mean, SD) | 1.2 (1.0) | 4.5 (1.9) | 1.7 (1.0) |
| **MET-hours per day age 52** (mean, SD) | 1.6 (0.9) | 4.4 (1.4) | 1.7 (1.1) |
| **MET-hours per day age 59** (mean, SD) | 1.2 (0.9) | 4.4 (2.1) | 3.0 (1.8) |
| **MET-hours per day age 91** (mean, SD) | 0.7 (0.7) | 1.7 (1.4) | 3.6 (1.8) |

Note. The characteristics between LTPA trajectories were compared with a $\chi^2$ test for sex (adjusted for clustered twin data) and with a non-parametrical Kruskal–Wallis test for education. Statistically significant differences between LTPA trajectories are marked with an asterisk. Abbreviations: LTPA, leisure-time physical activity; MET, metabolic equivalent of energy expenditure; SD, standard deviation.
\*p=0.009 (The trajectory *Starting high and decreasing* was significantly different from the *Constant low* and *Starting low and increasing* trajectories).

The second largest proportion of participants belonged to the LTPA trajectory labeled as *Starting low and increasing* and was characterized by a low level of LTPA at ages 45 and 52 with a clear increasing trend when approaching retirement age and the tenth decade of life. The mean leisure-time MET-hours were 1.7 (SD 1.0), 1.7 (SD 1.1), 3.0 (SD 1.8), and 3.6 (SD 1.8) at the mean ages of 45, 52, 59, and 91 years, respectively. In other words, the participants in this trajectory had a modest level of LTPA in their midlife but a moderate level in their sixties and nineties, approximately corresponding to a 30-minute daily walk at a slow pace and a 40-minute brisk daily walk, respectively.

The fewest participants were assigned to the *Starting high and decreasing* trajectory, which was characterized by the highest level of LTPA throughout the midlife years (i.e., working ages 45–59) with a decreasing trend from age 59 to age 91: the mean leisure-time MET-hours per day were 4.5 (SD 1.9), 4.4 (SD 1.4), 4.4 (SD 1.7), and 1.7 (SD 1.4) at the mean ages of 45, 52, 59 and 91 years, respectively. In practice, the participants in the *Starting high and decreasing* trajectory had a 1-hour brisk daily walk in their forties and fifties, while as nonagenarians, their LTPA corresponded to about a 30-minute daily walk at a slow pace.

There were significant differences in the educational level between the longitudinal LTPA trajectories identified. The study participants who belonged to the *Constant low* trajectory had the lowest level of education, while study participants belonging to the *Starting high and decreasing* trajectory had the highest education level on average. Our sensitivity analyses also indicated that the individuals who participated in the cognition assessments as nonagenarians were more educated than non-participants (p-value < 0.001) (see Supplemental Table S2).

### Longitudinal leisure-time physical activity trajectory associations with cognition

First, we examined whether participants’ cognition in each LTPA trajectory were different from cognition of the participants from other LTPA trajectories (one trajectory *versus* other trajectories). These analyses indicated no significant differences in cognition characteristics between trajectories (nominal p-values ≥ 0.30) when adjusted for age and sex. The second set of analyses dealt with the pairwise comparisons between trajectories (one trajectory *versus* another trajectory), and again, we found no significant differences in cognition characteristics between LTPA trajectories when adjusted for sex and age (nominal p-values ≥ 0.30). Both sets of analyses (i.e., (i) one trajectory *versus* other trajectories and (ii) one trajectory *versus* another trajectory) were conducted again with sex, age, and education treated as covariates. We found no significant differences in cognition in these fully adjusted analyses (nominal p-values ≥ 0.18 when one trajectory was compared to other trajectories and nominal p-values ≥ 0.261 when one trajectory was compared to another trajectory) (Figures 2 and 3, Supplemental Tables S3 – 6).

**Figure 2.**
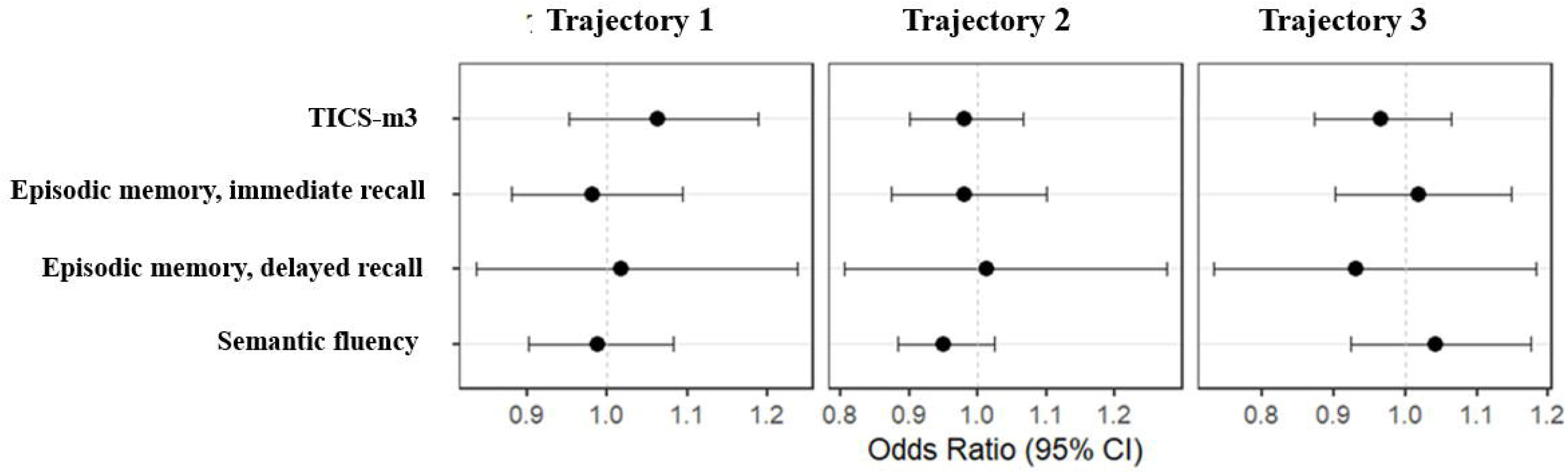
Summary statistics of cognition at age 91 by longitudinal leisure-time physical activity trajectories. Participants from one leisure-time physical activity trajectory were compared to all participants from other trajectories. Abbreviations: CI, confidence interval; TICS, Telephone Interview for Cognitive Status; Vs, versus.

**Figure 3.**
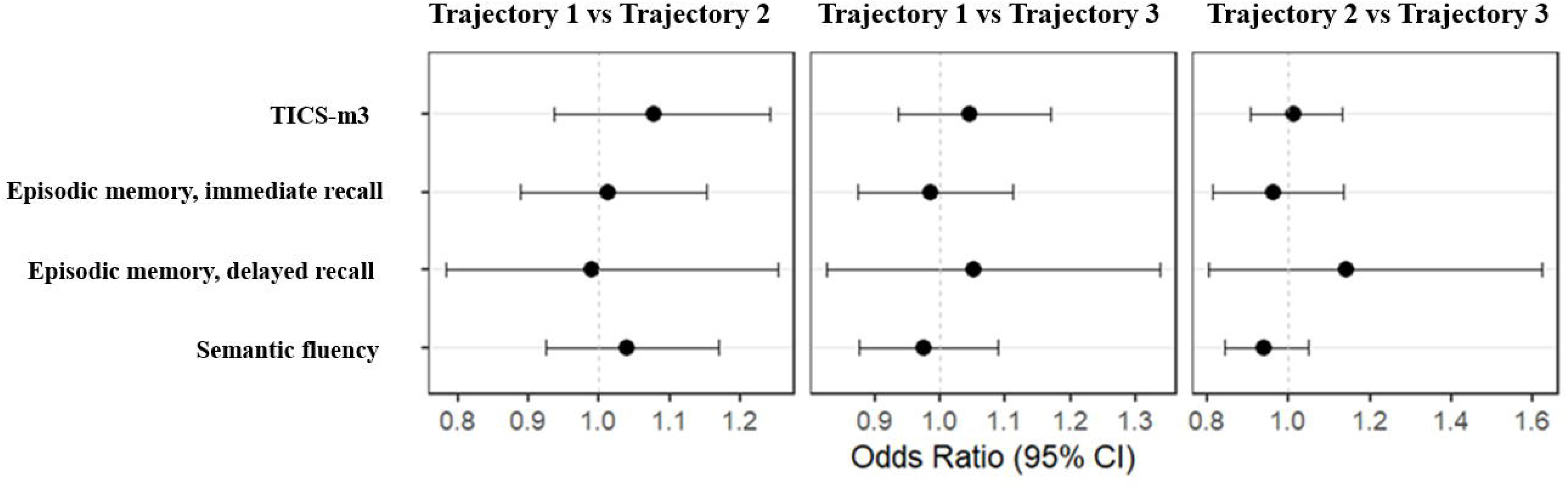
Summary statistics of pairwise comparisons of cognition at age 91 between each longitudinal leisure-time physical activity trajectory. Abbreviations: CI, confidence interval; TICS, Telephone Interview for Cognitive Status; Vs, versus.

### Longitudinal leisure-time physical activity trajectory associations with nonagenarians’ accelerometer-measured physical activity

The lowest LTPA trajectory (*Constant low*) was associated with the lowest mean number of steps as well as the lowest average times of light and moderate-to-vigorous physical activity per day at the nonagenarian age when sex and age were taken into account (odd ratios between 0.22–0.43, nominal p-values < 0.048). Nonagenarians’ accelerometer-measured mean number of steps and the average times of light and moderate-to-vigorous physical activity per day in each trajectory are shown in Table 2.

**Table 2.** The associations between longitudinal leisure-time physical activity trajectories and nonagenarians’ accelerometer-measured physical activity.

|  |  | Nonagenarians' accelerometer-measured physical activity |  |  |
| --- | --- | --- | --- | --- |
|  |  | Mean low PA time<br>per day<br>hh:mm:ss (SD) | Mean moderate-to-vigorous PA time<br>per day<br>hh:mm:ss (SD) | Mean number of<br>steps per day (SD) |
| **LTPA**<br>**trajecto-**<br>**ries** | **Constant Low** | **1:19:33\* (0:44:39)** | **0:05:33\* (0:05:46)** | **2030\* (1556)** |
|  | **Starting Low**<br>**and Increasing** | **2:06:27 (0:45:10)** | **0:17:28 (0:20:44)** | **4183 (3133)** |
|  | **Starting High**<br>**and Decreasing** | **1:49:16 (0:44:22)** | **0:15:07 (0:19:41)** | **3495 (2761)** |

Note. Statistically significant differences between LTPA trajectories are marked with an asterisk (The trajectory *Constant Low* was significantly different from the other two trajectories). Abbreviations: h, hour; LTPA, leisure-time physical activity; m, minute; PA, physical activity; s, second; SD, standard deviation.

## Discussion

To our knowledge, our study is the first to generate nearly-five-decades-long LTPA trajectories from midlife to the tenth decade of life. We found three different LTPA trajectories. The *Constant low* trajectory was characterized by the low stable level of LTPA throughout the follow-up from midlife to the mean age of 91. The *Starting low and increasing* trajectory was also characterized by low levels of LTPA in midlife but the participants in this trajectory first increased their LTPA level when they approached retirement age and as nonagenarians. The third LTPA trajectory labeled *Starting high and decreasing* portrayed a group of working age (ages 45, 52, and 59) individuals with moderate LTPA levels, but whose LTPA level notably decreased by the age of 91. Our results revealed that the longitudinal LTPA trajectory that was at the lowest level over the follow-up period was associated with the lowest number of steps as well as the lowest levels of light and moderate-to-vigorous physical activity measured with accelerometers in nonagenarians. The participants in *Constant low* trajectory also had significantly lower level of education than the participants from the other two trajectories.

Previous studies on the association between longitudinal physical activity trajectories and cognition have used various methods to identify longitudinal trajectories of physical activity, including growth mixture modeling,^8,10^ mixed models,^5^ latent process mixed models,^7^ and group-based trajectory modeling.^6,9,11–12^ The longitudinal physical activity trajectories found in these studies are very diverse and the number of trajectories varies from 2 to 5. A common feature for almost all studies presenting trajectories for absolute physical activity level is that they find one physical activity trajectory with a low stable pattern, and the largest proportion of participants belong to this trajectory.^6,9,12^ Of eight studies identifying physical activity trajectories and examining their associations with cognition, three have found a physical activity trajectory with a constant high^6,10,12^ and three with inconsistently high physical activity levels.^8,9,11^ Two studies have used a case–control approach, showing either lower physical activity levels in cases throughout the whole follow-up^7^ or diverging physical activity trajectories approximately nine years before dementia diagnosis between cases and controls.^5^ Our current results are consistent with these earlier studies, as we found one large constantly low physical activity trajectory and other physical activity trajectories had inconsistent physical activity levels. Our data-driven trajectory modeling provided us with one LTPA trajectory in which the highest LTPA levels were reached at ages 59 and 91. Similar increasing physical activity trajectories to ours have also been found in a few studies using model-based trajectory analysis methods.^6,8^

Perhaps our most important finding is that no significant association between nearly-five-decades-long LTPA trajectories and cognition in nonagenarians was found when age, sex, and education were considered. An association was not even found among the participants of the *Starting high and decreasing* trajectory, despite some previous studies having found that the decreasing physical activity trajectories seem to precede cognitive decline^8^ and dementia.^5^ On the other hand, our finding is consistent with earlier studies on physical activity and cognition in nonagenarians showing no association between these two.^15–17^ Most earlier studies have also found either low physical activity trajectories or decreasing physical activity trajectories preceding cognitive decline,^5–10^ but these studies have assessed cognition mostly in individuals aged 65–80 years. Thus, it seems that a relationship between physical activity and cognition may be stronger earlier in life and weakens in older adults aged 85 years and over. We have looked at the association of physical activity separately at each time point of 1975, 1981, 1990, and at 90 years old, in association with cognition at nonagenarian age. In line with our current trajectory approach, these results indicated that physical activity measured in midlife or in old age was not associated with cognition at 90 years old (Varjonen et al. 2025).^30^ Our finding is in accordance with the physical activity trajectory and cognition study from Gong et al.^11^ who similarly found that aging attenuates the positive association between physical activity trajectories and cognition. Our earlier meta-analysis showed no moderation by age on the association between physical activity and cognition, but we should be cautious about the result since the proportion of studies with nonagenarians included in the meta-analysis was small.^2^ Our results suggest that factors such as genes and education, for example, may be more influential in determining cognition than longitudinal LTPA behavior in exceptionally long-lived nonagenarians. This finding is in line with our earlier study showing a weak association between cardiovascular risk factors and cognition in nonagenarians but indicating that higher education is associated with better cognition even when controlling for other risk factors and with other studies showing no association between cardiovascular risk factors and dementia among older adults aged 85 years and over.^31–32^ Previous studies have revealed that the cognitive performance of individuals aged 85 years and over has improved in more recent age cohorts compared to older age cohorts.^33–34^ When researchers have considered reasons for this shift, they have not implicated improvement in lifestyles but rather the generally improved medical care (e.g., more intensive blood pressure treatment, lipid-lowering drugs, and antithrombotics)^34^ and the so-called Flynn effect (i.e., cohorts born later cognitively outperform cohorts born earlier).^33^ It may be that the uneven distribution of unmeasured factors (such as the quality of medical care, tendency to seek medical care or earlier general cognitive ability) or the general improvement in living standards from earlier times to current times contributes more for nonagenarian cognition than physical activity alone. At the same time, work-related physical activity has decreased substantially in the past five decades, which may represent an unmeasured confounder.

The different LTPA trajectories had significant differences in the length of education: the participants in the *Constant low* trajectory had significantly lower level of education. This finding that the participants with lowest physical activity trajectories have significantly lower level of education than participants from other trajectories is common.^5–6,9–12^ Earlier evidence shows that cognitive reserve, for which education serves as a proxy, moderates the association between lifestyles and cognitive aging so that cognitive reserve protects against deteriorative the effects of unfavorable life-styles.^35–36^ The lack of an association between LTPA trajectories and cognition in our nonagenarians may reflect this phenomenon: those who survive to nonagenarian age are selected in terms of education and in these individuals with higher level of education, the association between lifestyles and cognition is weaker.

The spectrum of neurodegenerative pathology in the brain differs according to age.^37^ In nonagenarians, limbic-predominant age-related TAR DNA-binding protein 43 (TDP-43) encephalopathy, brain arteriosclerosis, vascular dementia due to lacunar infarcts, large infarcts and microinfarcts, and primary age-related tauopathy explain a large proportion of neurodegeneration, while these pathologies are much rarer among individuals 60 to 80 years old.^37^ Alzheimer’s disease remains, however, the most prevailing neuropathology in both age groups. We have shown in our previous study that physical activity is associated with a lower incidence of both Alzheimer’s disease and vascular dementia.^3^ Consequently, the varying spectrum of neurodegeneration across aging is an unlikely explanation for the lack of association between 46-year LTPA trajectories and cognition in nonagenarians.

Mechanisms via which physical activity may decelerate both pathological and aging-related cognitive decline are numerous: cerebral blood flow, hippocampal volume, neuroinflammation, mitochondrial function, autophagy and proteasomal degradation, epigenetic modifications, brain plasticity, neurogenesis, and microbiota both in the gut and oral cavity.^38^ Despite this vast plethora of possible mechanisms via which physical activity may be beneficial for brain health, many studies show no gain despite the “pain”. For example, a meta-analysis of exercise intervention studies shows no influence on brain volume,^39^ and large longitudinal observational studies show a reverse association: brain volume predicts physical activity participation instead of vice versa.^40–41^ Even though super-agers tend to be both physically and mentally strong,^14^ the conundrum of which comes first, the chicken or the egg, better cognition or physical activity, continues. It must also be remembered that understanding aging is not a simple matter and associations between lifestyles and diseases, such as neurodegenerative conditions, are intertwined with many factors. For example, genetics are shown to affect both physical activity behavior^42^ and cognition.^43^

There are some limitations in our study that need to be considered. Although we constructed unique and exceptionally long LTPA trajectories, the greatest weakness of our study is the small sample size that may inhibit us from detecting a weak association between LTPA trajectories and cognition. However, a 46-year follow-up study with several health and behavior data from midlife to the tenth decade of life is a rare study design, and no very large sample size can be expected. Moreover, the follow-up time between the last two time points was 30 years: shorter intervals between LTPA assessments would have provided more precise data on LTPA patterns. It is also possible that our LTPA results are biased, given the self-reported nature of LTPA. However, the self-reported LTPA items we used have been shown to have high validity with other LTPA questionnaires in the Older Finnish Twin Cohort^21–22^ and accelerometer-measured physical activity behavior in the NONAGINTA study.^20^ Finally, cognition was assessed with a telephone interview in the present study. The telephone interview is more vulnerable to interference by hearing problems. Individuals with mild-to moderate hearing problems have stated, however, that they heard better via telephone than face-to-face and an earlier study of this cohort has shown that the overall performance in telephone cognition interview did not differ significantly between those using and not using hearing aid.^44^

The participants of the Older Finnish Twin Cohort have also previously participated in a cognition interview when they were 72 years old on average. Then, the participants of the cognition interview were more educated than non-participants (Iso-Markku et al. 2021). The participants of the current NONAGINTA sub-study (participants drawn from the Older Finnish Twin Cohort) with information on LTPA from at least two time points across the 46-year follow-up are even more educated (8.4 years of education vs 8.0 at mean age 72^45^). Therefore, our nonagenarian participants are selected in terms of education (and likely other attributes as well). Compared to many study cohorts, the formal education level in the population-based Older Finnish Twin Cohort is, however, still exceptionally low and unselected as compulsory education ended after six years in these birth cohorts. Only a small fraction of school children would go on to secondary or tertiary education in the early post-war years after the Continuation War (1941-1944). In addition, mortality and dementia share risk factors like APOE genotype,^46–47^ and education.^48–49^ Thus, survivorship bias may also hinder us from detecting LTPA-induced differences in nonagenarians’ cognition.

The main strength of this study is the 46-year follow-up of LTPA. As far as we know, our current study examining prior lifestyle habits extending from age 45 to almost 5 decades ahead is unprecedented. Overall, studies on the determinants of nonagenarians’ cognition are also very rare. We also assessed LTPA with many survey questions, allowing for more precise estimates of LTPA levels: the frequency, intensity and duration of LTPA, including active commuting, were assessed at each follow-up time point, except at the last follow-up time point when participants were 91-year-olds on average and commuting activity was no longer relevant. An additional strength is that our population-based study cohort has a quite equal sex representation. Thus, our results may be more generalizable at the population level.

In conclusion, we have provided new knowledge on longitudinal LTPA trajectories from midlife to the tenth decade of life and their association with nonagenarian cognition. Nearly-five-decades-long LTPA trajectories were not significantly associated with nonagenarians’ cognition. Based on our study, other factors, such as education, are likely to be more important in determining cognition in nonagenarians. Our results may also be partly explained by survivorship bias and a small sample size. Studies with a larger sample size on the topic could provide more definitive evidence in the future and are, thus, warranted. Moreover, as the number of nonagenarians continues to rise, more research should be directed to disentangling potential factors affecting cognition in older adults aged 85 years and over. Although we did not find significant associations between nearly-five-decades-long LTPA trajectories and nonagenarians’ cognition, LTPA is recommendable due to its other numerous benefits for cardiovascular function, physical function, and mental health.^50^

## Data Availability

The Older Finnish Twin Cohort data are not publicly available due to the restrictions of informed consent. However, the data are available through the Institute for Molecular Medicine Finland (FIMM) Data Access Committee (DAC) for authorized researchers who have IRB/ethics approval and an institutionally approved study plan. To ensure the protection of privacy and compliance with national data protection legislation, a data use/transfer agreement is needed, the content and specific clauses of which will depend on the nature of the requested data.

## Conflicts of interest

None.

## Funding

This work was supported by the Academy of Finland (grants 314639, 320109 and 345988 to E.V.), Päivikki and Sakari Sohlberg Foundation (S.A. and P.I.M.), the Finnish Cultural Foundation (S.A. and P.I.M.), Juho Vainio Foundation (S.A.), Orion Research Foundation (P.I.M.), Biomedicum Research Foundation (P.I.M.), and the Sigrid Jusélius Foundation (E.V.). The funders had no role in study design, data collection and analysis, decision to publish, or preparation of the manuscript. Jaakko Kaprio acknowledges support from the Academy of Finland Center of Excellence in Complex Disease Genetics (grants 336823 & 352792).

## Acknowledgements

We thank Kauko Heikkilä and Teemu Palviainen for the data management, and Mia Urjansson for the cognition assessments among nonagenarians. We are especially grateful for all the twins who have participated in the Finnish Twin Cohort study enabling this investigation.

## Author contributions

J.K., E.V., S.A., H.V., and T.V. acquired the clinical data. S.A., J.K., E.V., G.D., V.F., and P.I. conceived and designed the study. S.A., P.I., G.D., V.F., H.V., and A.V. processed and analyzed the data, and all authors contributed into the interpretation of the data. P.I., S.A., and G.D. drafted the manuscript. P.I. and G.D. made the visualizations. All authors provided intellectual input to the manuscript and critically revised the manuscript. All authors have read and approved the final version of the manuscript.

## Notes

### Competing Interest Statement

The authors have declared no competing interest.

### Funding Statement

This work was supported by the Academy of Finland (grants 314639, 320109 and 345988 to E.V.), Paivikki and Sakari Sohlberg Foundation (S.A. and P.I.M.), the Finnish Cultural Foundation (S.A. and P.I.M.), Juho Vainio Foundation (S.A.), Orion Research Foundation (P.I.M.), Biomedicum Research Foundation (P.I.M.), and the Sigrid Juselius Foundation (E.V.). The funders had no role in study design, data collection and analysis, decision to publish, or preparation of the manuscript. Jaakko Kaprio acknowledges support from the Academy of Finland Center of Excellence in Complex Disease Genetics (grants 336823 & 352792).

### Author Declarations

The data collection was approved by the ethics committees of the Hjelt Institute, University of Helsinki and the Helsinki and Uusimaa Hospital District, Finland. The latest ethical approval was given by the Ethics Committee of the Hospital District of Helsinki and Uusimaa for the NONAGINTA study protocol on May 8, 2020 and December 16, 2020 (the latest follow-up time point in the current study).

