## Supplemental file for "Data-Driven Leisure-Time Physical Activity Trajectories of Over 46 Years and Their Associations with Cognition in Nonagenarians – A Cohort Study"

This PDF file includes:

Supplemental text

Supplemental Figures S1 – 3

Supplemental Tables S1 – 6

Leisure-time physical activity questions at ages 45, 52, 59, and 91

### 1 Supplemental Text

#### 1.1 Materials and methods

##### 1.1.1 Self-reported physical activity

To calculate MET hours per day from the LTPA questions, we used the following formula: LTPA frequency (average per day) × LTPA duration (average hours) × LTPA intensity (activity MET score) (+ average active commuting per day at mean ages 45, 52, 59). The following MET score values were used for the intensity of LTPA to obtain a multiple of the resting metabolic rate for each activity: 4 corresponded to walking, 6 corresponded to vigorous walking to jogging, 10 corresponded to jogging, and 13 corresponded to running. The MET value of 4 (walking) was also used for the intensity of commuting-related physical activity. We further assumed that commuting-related physical activity was done five days per week. All types of LTPAs were considered when MET hours per day were calculated.

##### 1.1.2 Accelerometer-measured physical activity

Hip-worn tri-axial accelerometer (UKK RM42, UKK Terveyspalvelut Oy, Tampere, Finland) along with instructions were mailed to the study participants of the telephone interview of NONAGINTA study. The accelerometers were advised to be worn on right side of the hip with an elastic band for seven consecutive days during waking hours except for shower, swimming and bathing. Measurements with at least 10 hours wear-time per day and worn on at least 4 days were included in the analyses. The acceleration data were recorded at a sampling frequency of 100 Hz. We used mean amplitude deviation (MAD) and angle for posture estimation (APE) algorithms in the analyses when calculating MET-hours from the raw data. MET-values were smoothed by calculating the moving average for each 6-second epoch.

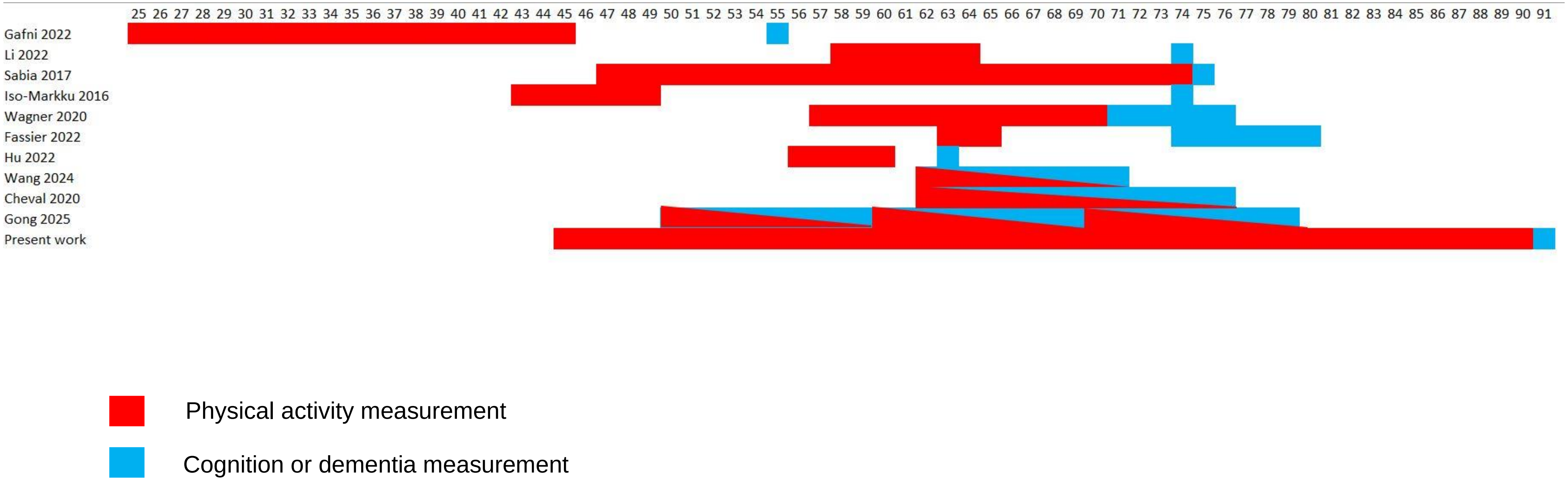


* Time is presented as years of age.

Supplemental Figure S1. Timeline of observational studies assessing the association of longitudinal physical activity (measured at multiple time points) and cognition

**Supplemental Table S1.** The comparison of six different clustering solutions in identifying 46-year-long leisure-time physical activity trajectories using K-mean cluster modeling.

| Number of clusters | Calinski-Haribasz index | Calinski-Harabasz index 2 | Calinski-Harabasz index 3 | Ray-Turi | Davies-Bouldin | Sizes of clusters |
|---|---|---|---|---|---|---|
| 2 | 78.15186910 | 0.6405467492 | 78.15186910 | -0.0887908130 | -1.276220481 | 82;43 |
| **3** | **69.14427808** | **1.152094931** | **97.78477582** | **-0.1085795173** | **-1.338537211** | **65;31;29** |
| 3 | 69.16918789 | 1.152509983 | 97.82000361 | -0.1105532049 | -1.341208561 | 65;33;37 |
| 4 | 58.53578732 | 1.487283169 | 101.3869577 | -0.1490310090 | -1.370455004 | 65;26;25;13 |
| 5 | 52.23451244 | 1.799188761 | 104.4690248 | -0.2823481970 | -1.399690875 | 40;29;25;19;12 |
| 6 | 49.3854007 | 2.162202418 | 110.4291131 | -0.2786909543 | -1.457110478 | 39;29;18;15;12;12 |

Note. The final solution selected is in bold.

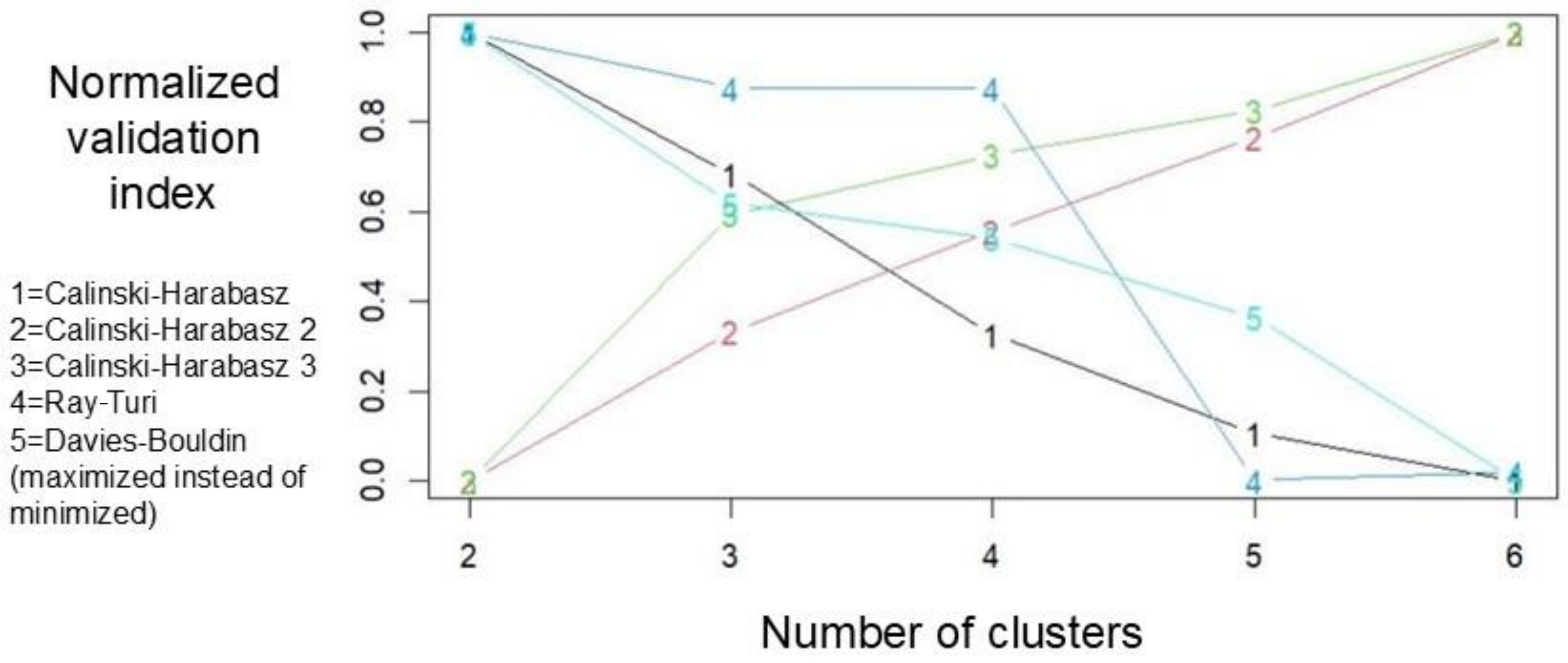


Supplemental Figure S2. Validation indices in identifying leisure-time physical activity trajectories using K-mean cluster modeling

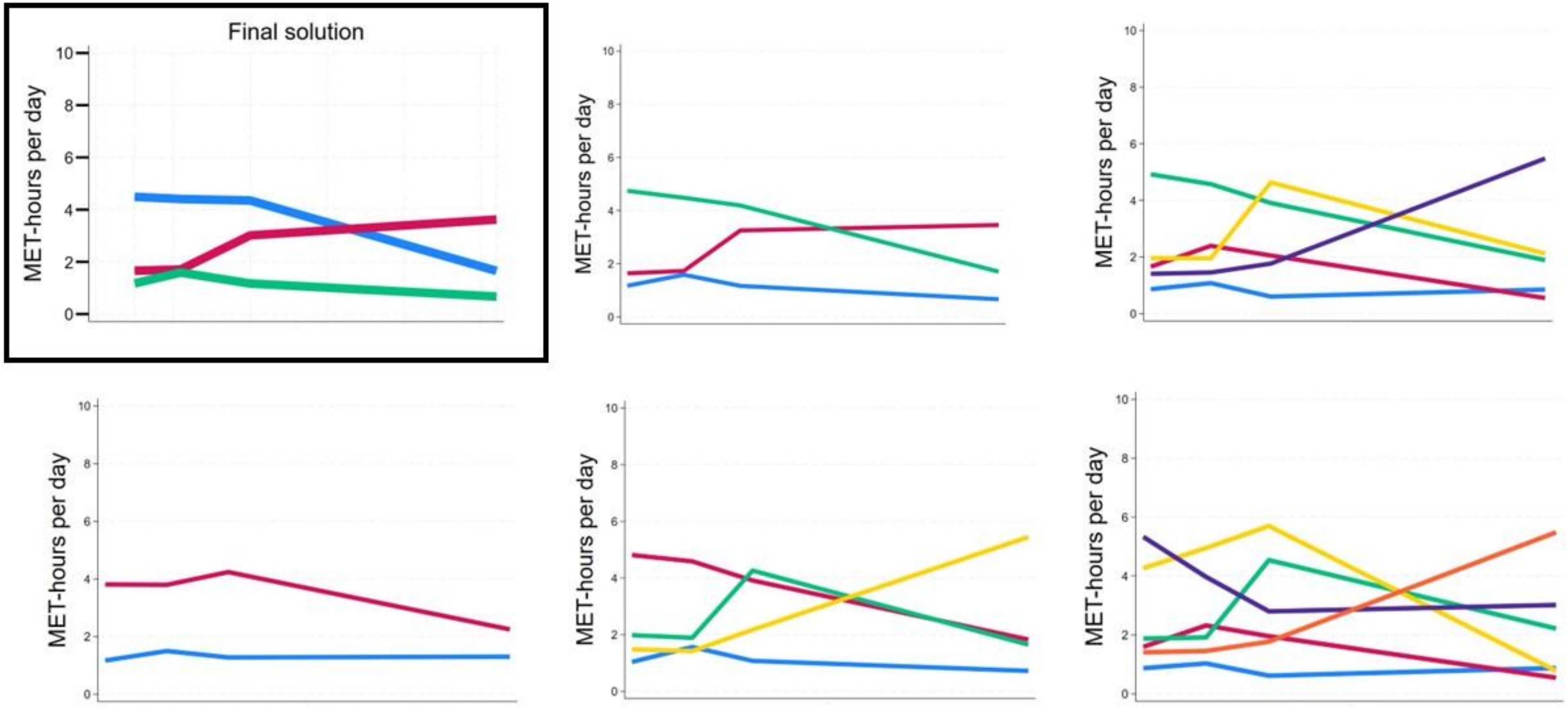


Supplemental Figure S3. Six examples of clustering solutions in identifying leisure-time physical activity trajectories using K-mean cluster modeling

**Supplemental Table S2.** The comparison of study participants' characteristics between the participants and non-participants of cognitionassessments

| | **Participated in cognition assessment** (n=80) | **Did not participate in cognition assessment** (n=45) | **P-value for difference between groups** * |
|---|---|---|---|
| **Sex** | | | |
| Men(n, %) | 33 (41.3) | 14 (29.2) | |
| Women(n, %) | 47 (58.5) | 34 (70.8) | 0.093 |
| **Education years** (mean, SD) | 9.4 (4.6) | 6.6 (1.6) | < 0.001 |
| **MET-hours per day age 45** (mean, SD) | 2.2 (1.9) | 1.9 (1.8) | 0.388 |
| **MET-hours per day age 52** (mean, SD) | 2.5 (1.7) | 2.0 (1.4) | 0.165 |
| **MET-hours per day age 59** (mean, SD) | 2.6 (2.1) | 2.2 (1.6) | 0.384 |
| **MET-hours per day age 91** (mean, SD) | 1.7 (1.6) | 1.6 (1.9) | 0.849 |

* Difference between groups has been compared with t-test (adjusted for clustered twin data)

Abbreviations: MET, metabolic equivalent of energy expenditure; SD, standard deviation.

**Supplemental Table S3.**Differences in cognition by longitudinal LTPAtrajectories. Participants from one LTPA trajectory were compared to all participants from other trajectories. The analyses were adjusted for age and sex.

| | | | | | Confidence intervals | | p-values | |
|---|---|---|---|---|---|---|---|---|
| **Variable** | **Trajectory class** | **Mean, trajectory class** | **Mean, other trajectory classes** | **Odds ratio** | **Lower bound** | **Upper bound** | **Nominal** | **Benjamini–Hochberg** |
| Semanticfluency | 1 | 14.56 | 15.59 | 0.96 | 0.87 | 1.04 | 0.313 | 0.907 |
| Semanticfluency | 2 | 14.69 | 15.16 | 0.98 | 0.91 | 1.07 | 0.681 | 0.907 |
| Semanticfluency | 3 | 16.22 | 14.60 | 1.06 | 0.95 | 1.20 | 0.296 | 0.907 |
| TICS-m3 | 1 | 37.07 | 39.33 | 0.98 | 0.92 | 1.04 | 0.454 | 0.907 |
| TICS-m3 | 2 | 39.07 | 38.04 | 1.01 | 0.95 | 1.08 | 0.723 | 0.907 |
| TICS-m3 | 3 | 39.53 | 37.72 | 1.01 | 0.94 | 1.08 | 0.786 | 0.907 |
| Episodic memory, immediate recall | 1 | 11.37 | 12.21 | 0.93 | 0.83 | 1.04 | 0.198 | 0.907 |
| Episodic memory, immediate recall | 2 | 11.94 | 11.73 | 1.02 | 0.91 | 1.14 | 0.756 | 0.907 |
| Episodic memory, immediate recall | 3 | 12.39 | 11.53 | 1.05 | 0.94 | 1.18 | 0.413 | 0.907 |
| Episodic memory, delayed recall | 1 | 2.24 | 2.56 | 0.94 | 0.78 | 1.14 | 0.534 | 0.907 |
| Episodic memory, delayed recall | 2 | 2.62 | 2.34 | 1.08 | 0.85 | 1.36 | 0.527 | 0.907 |
| Episodic memory, delayed recall | 3 | 2.52 | 2.35 | 1.00 | 0.82 | 1.23 | 0.975 | 0.993 |
| TICS-m | 1 | 28.10 | 29.48 | 0.98 | 0.91 | 1.06 | 0.663 | 0.907 |
| TICS-m | 2 | 29.43 | 28.67 | 1.02 | 0.92 | 1.12 | 0.720 | 0.907 |
| TICS-m | 3 | 29.53 | 28.53 | 1.00 | 0.91 | 1.10 | 0.993 | 0.993 |

Note. Trajectory class 1 = Constant Low; Trajectory class 2 = Starting Low and Increasing; Trajectory class 3 = Starting High and Decreasing; LTPA, leisure-time physical activity; TICS-m =the total score of TICS-m after one-word list learning trial; TICS-m3 = the total score of TICS-m after three-word list learning trial

**Supplemental Table S4.**Pairwise comparisons of cognition between each longitudinal LTPA trajectory. The analyses were adjusted for age and sex.

| | | | | | | Confidence intervals | | p-values | |
|---|---|---|---|---|---|---|---|---|---|
| **Variable** | **Trajectory class A** | **Trajectory class B** | **Mean, trajectory class A** | **Mean, trajectory class B** | **Odds ratio** | **Lower bound** | **Upper bound** | **Nominal** | **Benjamini–Hochberg** |
| TICS-m3 | 1 | 2 | 37.07 | 39.07 | 0.98 | 0.90 | 1.06 | 0.569 | 0.964 |
| Episodic memory, immediate recall | 1 | 2 | 11.37 | 11.94 | 0.96 | 0.84 | 1.09 | 0.481 | 0.964 |
| Episodic memory, delayed recall | 1 | 2 | 2.24 | 2.62 | 0.93 | 0.73 | 1.18 | 0.533 | 0.964 |
| Semantic fluency | 1 | 2 | 14.56 | 14.69 | 0.99 | 0.89 | 1.10 | 0.822 | 0.964 |
| TICS-m3 | 1 | 3 | 37.07 | 39.53 | 0.98 | 0.92 | 1.05 | 0.573 | 0.964 |
| Episodic memory, immediate recall | 1 | 3 | 11.37 | 12.39 | 0.95 | 0.84 | 1.07 | 0.366 | 0.964 |
| Episodic memory, delayed recall | 1 | 3 | 2.24 | 2.52 | 0.99 | 0.79 | 1.23 | 0.910 | 0.964 |
| Semantic fluency | 1 | 3 | 14.56 | 16.22 | 0.94 | 0.85 | 1.05 | 0.300 | 0.964 |
| TICS-m3 | 2 | 3 | 39.07 | 39.53 | 1.00 | 0.92 | 1.08 | 0.937 | 0.964 |
| Episodic memory, immediate recall | 2 | 3 | 11.94 | 12.39 | 0.97 | 0.84 | 1.12 | 0.646 | 0.964 |
| Episodic memory, delayed recall | 2 | 3 | 2.62 | 2.52 | 1.05 | 0.80 | 1.39 | 0.714 | 0.964 |
| Semantic fluency | 2 | 3 | 14.69 | 16.22 | 0.94 | 0.84 | 1.05 | 0.295 | 0.964 |
| TICS-m | 1 | 2 | 28.10 | 29.43 | 0.98 | 0.86 | 1.10 | 0.701 | 0.964 |
| TICS-m | 1 | 3 | 28.10 | 29.53 | 0.99 | 0.90 | 1.09 | 0.878 | 0.964 |
| TICS-m | 2 | 3 | 29.43 | 29.53 | 1.00 | 0.90 | 1.12 | 0.964 | 0.964 |

Note. Trajectory class 1 = Constant Low; Trajectory class 2 = Starting Low and Increasing; Trajectory class 3 = Starting High and Decreasing; LTPA, leisure-time physical activity; TICS-m =the total score of TICS-m after one-word list learning trial; TICS-m3 = the total score of TICS-m after three-word list learning trial.

**Supplemental Table S5.** Differences in cognition by longitudinal LTPA trajectory. Participants from one LTPA trajectorywere compared to all participants from other trajectory. The analyses were adjusted for age, sex, and education.

| | | | | | Confidence intervals | | p-values | |
|---|---|---|---|---|---|---|---|---|
| **Variable** | **Trajectory class** | **Mean, trajectory class** | **Mean, other trajectory classes** | **Odds ratio** | **Lower bound** | **Upper bound** | **Nominal** | **Benjamini–Hochberg** |
| TICS-m3 | 1 | 37.07 | 39.33 | 1.02 | 0.96 | 1.10 | 0.493 | 0.903 |
| TICS-m3 | 2 | 39.07 | 38.04 | 0.98 | 0.92 | 1.04 | 0.565 | 0.903 |
| TICS-m3 | 3 | 39.53 | 37.72 | 0.99 | 0.92 | 1.06 | 0.756 | 0.903 |
| Episodic memory, immediate recall | 1 | 11.37 | 12.21 | 0.98 | 0.88 | 1.09 | 0.737 | 0.903 |
| Episodic memory, immediate recall | 2 | 11.94 | 11.73 | 0.98 | 0.87 | 1.10 | 0.737 | 0.903 |
| Episodic memory, immediate recall | 3 | 12.39 | 11.53 | 1.02 | 0.90 | 1.15 | 0.776 | 0.903 |
| Episodic memory, delayed recall | 1 | 2.24 | 2.56 | 1.02 | 0.84 | 1.24 | 0.861 | 0.903 |
| Episodic memory, delayed recall | 2 | 2.62 | 2.34 | 1.01 | 0.81 | 1.28 | 0.903 | 0.903 |
| Episodic memory, delayed recall | 3 | 2.52 | 2.35 | 0.93 | 0.73 | 1.18 | 0.554 | 0.903 |
| Semantic fluency | 1 | 14.56 | 15.59 | 0.99 | 0.90 | 1.08 | 0.790 | 0.903 |
| Semantic fluency | 2 | 14.69 | 15.16 | 0.95 | 0.88 | 1.02 | 0.184 | 0.903 |
| Semantic fluency | 3 | 16.22 | 14.60 | 1.04 | 0.92 | 1.18 | 0.500 | 0.903 |
| TICS-m | 1 | 28.10 | 29.48 | 1.06 | 0.95 | 1.19 | 0.268 | 0.903 |
| TICS-m | 2 | 29.43 | 28.67 | 0.98 | 0.90 | 1.07 | 0.635 | 0.903 |
| TICS-m | 3 | 29.53 | 28.53 | 0.97 | 0.87 | 1.07 | 0.477 | 0.903 |

Note. Trajectory class 1 = Constant Low; Trajectory class 2 = Starting Low and Increasing; Trajectory class 3 = Starting High and Decreasing; LTPA, leisure-time physical activity; TICS-m =the total score of TICS-m after one-word list learning trial; TICS-m3 = the total score of TICS-m after three-word list learning trial

**Supplemental Table S6.** Pairwise comparisons of cognition between each longitudinal LTPA trajectory. The analyses were adjusted for age, sex, and education.

| | | | | | | Confidence intervals | | p-values | |
|---|---|---|---|---|---|---|---|---|---|
| **Variable** | **Trajectory class A** | **Trajectory class B** | **Mean, trajectory class A** | **Mean, trajectory class B** | **Odds ratio** | **Lower bound** | **Upper bound** | **Nominal** | **Benjamini–Hochberg** |
| TICS-m3 | 1 | 2 | 37.07 | 39.07 | 1.03 | 0.95 | 1.13 | 0.439 | 0.993 |
| Episodic memory, immediate recall | 1 | 2 | 11.37 | 11.94 | 1.01 | 0.89 | 1.15 | 0.860 | 0.993 |
| Episodic memory, delayed recall | 1 | 2 | 2.24 | 2.62 | 0.99 | 0.78 | 1.25 | 0.936 | 0.994 |
| Semantic fluency | 1 | 2 | 14.56 | 14.69 | 1.04 | 0.92 | 1.17 | 0.519 | 0.993 |
| TICS-m3 | 1 | 3 | 37.07 | 39.53 | 1.01 | 0.94 | 1.09 | 0.734 | 0.993 |
| Episodic memory, immediate recall | 1 | 3 | 11.37 | 12.39 | 0.99 | 0.87 | 1.11 | 0.807 | 0.993 |
| Episodic memory, delayed recall | 1 | 3 | 2.24 | 2.52 | 1.05 | 0.83 | 1.34 | 0.684 | 0.993 |
| Semantic fluency | 1 | 3 | 14.56 | 16.22 | 0.98 | 0.88 | 1.09 | 0.662 | 0.993 |
| TICS-m3 | 2 | 3 | 39.07 | 39.53 | 1.00 | 0.91 | 1.10 | 0.993 | 0.994 |
| Episodic memory, immediate recall | 2 | 3 | 11.94 | 12.39 | 0.96 | 0.81 | 1.13 | 0.634 | 0.993 |
| Episodic memory, delayed recall | 2 | 3 | 2.62 | 2.52 | 1.14 | 0.80 | 1.62 | 0.462 | 0.993 |
| Semantic fluency | 2 | 3 | 14.69 | 16.22 | 0.94 | 0.84 | 1.05 | 0.261 | 0.993 |
| TICS-m | 1 | 2 | 28.10 | 29.43 | 1.08 | 0.94 | 1.24 | 0.295 | 0.993 |
| TICS-m | 1 | 3 | 28.10 | 29.53 | 1.05 | 0.94 | 1.17 | 0.428 | 0.993 |
| TICS-m | 2 | 3 | 29.43 | 29.53 | 1.01 | 0.90 | 1.13 | 0.847 | 0.993 |

Note. Trajectory class 1 = Constant Low; Trajectory class 2 = Starting Low and Increasing; Trajectory class 3 = Starting High and Decreasing; LTPA; leisure-time physical activity; TICS-m =the total score of TICS-m after one-word list learning trial; TICS-m3 = the total score of TICS-m after three-word list learning trial.

### Leisure-time physical activity questions

Questions have been translated from Finnish.

**Leisure-time physical activity question at the mean ages 45 and 52:**

How much of your daily journey to work/study is spent in walking, cycling, running and/or cross-country skiing?
1 less than 15 min
2 15 min to less than half an hour
3 half an hour to less than one hour
4 one hour or more
5 I am presently not at work/studying

How often do you exercise/engage in physical activity during your leisure time?
1 less than once a month
2 1–2 times a month
3 3–5 times a month
4 6–10 times a month
5 11–19 times a month
6 more than 20 times a month

Is your physical activity during leisure time about as tiring on average as:
1 walking
2 alternatively walking and jogging
3 jogging (light run)
4 running

How long does one session of the physical activity last on average?
1 less than 15 min
2 15 min to less than half an hour
3 half an hour to less than one hour
4 one hour to under two hours
5 two hours or more

**Leisure-time physical activity question at mean age 59:**

In the next question, we are going to inquire about your leisure-time and commuting physical activity during the last 12 months. We have divided physical activity into four different intensities. First, evaluate how strenuous each of the physical activities you engage in are. Then, evaluate, on average, how many hours per week you engage in physical activity corresponding to each intensity level?

| Intensity level of each physical activity | Not at all | Altogether less than ½ hour per week | Altogether ½ – 1 hour per week | Altogether 2 – 3 hours per week | Altogether 4 hours or more per week |
|---|---|---|---|---|---|
| Walking | 1 | 2 | 3 | 4 | 5 |
| Alternatively walking and jogging | 1 | 2 | 3 | 4 | 5 |
| Jogging (light run) | 1 | 2 | 3 | 4 | 5 |
| Running | 1 | 2 | 3 | 4 | 5 |

**Leisure-time physical activity questions at mean age 91:**

How often do you exercise/engage in physical activity sessions?

1 less than once a month
2 1–2 times a month
3 3–5 times a month
4 6–10 times a month
5 11–19 times a month
6 more than 20 times a month

Is your physical activity about as tiring on average as:
1 walking
2 alternatively walking and jogging
3 jogging (light run)
4 running

How long does one session of the physical activity last on average?
1 less than 15 min
2 15 min to less than half an hour
3 half an hour to less than one hour
4 one hour to under two hours
5 two hours or more
